# Whole Genome HPV Liquid Biopsy for Pan-HPV-Associated Cancer Detection and Viral Physical State Classification

**DOI:** 10.64898/2026.04.27.26350528

**Authors:** Adam S. Fisch, Annie R. Abruzzo, Samuli Eldfors, Dipon Das, Qin Wang, Gjystina Lumaj, Shriya Shukla, Allison A. Gockley, Jennifer Y. Wo, Theodore S. Hong, Andrea L. Russo, Jeremy D. Richmon, Fantine Giap, Bayan A. Alzumaili, William C. Faquin, Peter M. Sadow, Daniel L. Faden

**Affiliations:** Departments of Pathology, Massachusetts General Hospital; Harvard Medical School; Department of Otolaryngology-Head and Neck Surgery, Mass Eye and Ear; Division of Gynecologic Oncology, Department of Obstetrics, Gynecology, and Reproductive Biology, Massachusetts General Hospital, Boston, MA, USA; Department of Radiation Oncology, Massachusetts General Hospital, Boston, Massachusetts; Broad Institute of MIT and Harvard, Cambridge, MA, USA

## Abstract

**Purpose:** HPV-associated carcinomas (HPV+ cancers) account for 5% of all cancers. Circulating tumor HPV DNA (ctHPVDNA) assays for HPV+ cancer surveillance have limited prognostic utility at the time of cancer diagnosis. While HPV integration into the host genome is a proven tissue-based biomarker predicting poor clinical outcomes, existing clinically utilized ctHPVDNA assays cannot classify the viral physical state.

**Methods:** We previously developed HPV-DeepSeek, a multi-feature HPV whole-genome sequencing liquid biopsy with 99% diagnostic accuracy at the time of HPV+ oropharynx cancer diagnosis. We test the diagnostic accuracy of HPV-DeepSeek in a cohort of 235 HPV+ cancers across nine anatomic sites and employ a novel blood-based computational classifier to infer HPV genome physical state from plasma, termed HPV-SIGNAL, to assess its prognostic potential.

**Results:** HPV-DeepSeek demonstrated a sensitivity and specificity of 99%. In 181 eligible samples, HPV-SIGNAL identified four viral physical states: episomal-only (N = 69), episomal-rearranged (N = 48), integrated-mixed (N = 55), and integrated-clonal (N = 9), which were confirmed and further elucidated via three orthogonal tissue and blood approaches. Patients harboring integrated viral states in the blood exhibited significantly worse progression-free survival (HR 3.28, 95% CI 1.63-6.58, p = 0.00084) and overall survival (HR 2.98, 95% CI 1.16-7.64, p = 0.023) compared to patients with episomal states.

**Conclusion:** HPV whole-genome sequencing liquid biopsy has high diagnostic accuracy across HPV+ cancer types and can be used to identify and classify HPV physical state from blood. Patients with integrated viral states detected in the blood demonstrated worse progression-free and overall survival, suggesting blood-based HPV physical state classification could be used as a prognostic tool at the time of cancer diagnosis.

**Translational Relevance:** Current circulating tumor HPV DNA assays for HPV-associated cancer surveillance have limited prognostic utility at the time of cancer diagnosis. While HPV integration into the host genome is a proven tissue-based biomarker predicting poor clinical outcomes, existing circulating tumor HPV DNA assays cannot classify the viral physical state. Here, we show that HPV-SIGNAL, a novel blood-based computational classifier to infer HPV genome physical state from plasma using output from HPV-DeepSeek, an HPV whole genome sequencing liquid biopsy, accurately identifies and classifies HPV physical state from blood and is prognostic of progression-free and overall survival across HPV-associated cancer types.

## Introduction

Together, HPV-associated cancers cause 5% of all cancers worldwide^1–4^. Detection of circulating tumor HPV DNA (ctHPVDNA) is in routine clinical use in HPV-associated oropharyngeal cancer, the most common HPV-associated cancer, as a surveillance tool, and there is growing evidence in HPV-associated cervical cancer and anal cancer, the second and third most common HPV-associated types, respectively^3,5–23^. Current clinically active approaches for ctHPVDNA detection utilize droplet digital PCR (ddPCR); however, sensitivity is limited, with < 90% of patients having detectable ctHPVDNA at the time of cancer diagnosis and only 40% of patients having recurrence detected earlier than standard care^5^. Furthermore, current approaches cannot annotate the myriad of known prognostic factors in HPV-associated cancers, most notably viral physical state, which has been shown in tissue-based studies to predict recurrence and survival across multiple HPV-associated cancer types^24,25,25–27^.

We have previously described a multi-feature HPV whole genome sequencing liquid biopsy-based assay, HPV-DeepSeek, and demonstrated its high diagnostic accuracy at the time of diagnosis with HPV+ oropharynx cancer (0.99), in HPV+ oropharynx cancer screening and MRD detection^7,10,28^. Here, we describe and orthogonally validate a novel blood-based viral physical state computational classifier for HPV-DeepSeek, HPV-SIGNAL, and apply this across a multitude of anatomic sites representing the diversity of HPV-associated cancers, finding that the HPV physical state detection in plasma is prognostic for recurrence and survival across HPV-associated cancers.

## Materials and Methods

All participants of the study provided written informed consent to a protocol approved by the Dana Farber/Harvard Cancer Center (DFHCC 18-653). This study was conducted in compliance with the U.S. Common Rule. Blood samples were collected from 235 patients with HPV+ cancer prior to any cancer treatment. All patients underwent standard of care diagnostic workup. Study controls were enrolled at Mass General Brigham and composed of a mix of healthy individuals and non-HPV cancer controls presenting for health care. All blood samples were assigned a study ID and blinded for further analysis.

## Plasma DNA Extraction and Sequencing for HPV-DeepSeek

Cell-free DNA (cfDNA) extraction, sequencing, and analysis were performed as described previously^7^. Briefly, cfDNA was extracted from 1-4 mL of frozen plasma, and 20 ng cfDNA was used as input for hybrid-capture library preparation using the KAPA HyperCap cfDNA Workflow v1.0 (Roche) with UMI adapters and indexed amplification, with library quantification on Qubit (Thermo Fisher Scientific) and sizing/QC on TapeStation (Agilent). Target enrichment used customized KAPA HyperCap probes and the HyperCapture bead kit (Roche), followed by paired-end sequencing on an Illumina platform. Sequencing reads were adapter-trimmed and quality-filtered prior to alignment. Reads were mapped to a combined reference consisting of the human genome (hg38) and HPV reference sequences using BWA-MEM (v0.7.17)^29^, followed by UMI deduplication to remove PCR duplicates. HPV read counts were calculated from deduplicated alignments, and HPV genome coverage was defined as the percentage of positions in the HPV reference genome with at least one mapped read. HPV16 sublineage assignments were derived from the alignment data as previously described^7^.

## HPV Genome Physical State Classification with HPV-SIGNAL

We developed a classifier to infer HPV genome physical state from HPV-DeepSeek targeted short-read sequencing of ctHPVDNA, termed HPV Sequencing-based INteGration AnaLysis (HPV-SIGNAL). For each sample, we identified the HPV type with the highest HPV read count and classified physical state for this dominant HPV type. The classifier combined evidence from viral-human junctions, extent of HPV genome coverage, and HPV-HPV structural rearrangements to assign mutually exclusive molecular classes. Application of the classifier was restricted to samples meeting a minimum HPV signal threshold with classification of the dominant HPV type requiring at least 500 HPV-mapped reads after UMI deduplication for eligibility. UMI-deduplicated alignment files were screened for chimeric split reads identified by supplementary alignments (SA tag) with MAPQ ≥ 20. Breakpoint coordinates were inferred from CIGAR strings. Candidate breakpoint calls were excluded if the HPV breakpoint mapped to a terminus of the linear HPV reference sequence, as circular HPV genomes can generate artifactual breakpoint calls at reference ends. HPV-human integration and HPV-HPV rearrangement breakpoint calls were identified from chimeric reads, requiring at least 6 supporting reads per breakpoint call. Junctions were defined by clustering breakpoint calls based on HPV coordinate, with calls within 10 bp merged into a single junction event.

The inferred HPV physical state for each sample was assigned to 1 of 4 mutually exclusive classes using HPV-human junctions, HPV-HPV junctions, and HPV genome coverage. Near-complete HPV genome coverage was defined as coverage of at least 97% of positions in the HPV reference genome: 1) Episomal-only samples had complete/near-complete HPV genome coverage and no detected junctions, consistent with intact full-length episomal HPV DNA. 2)

Episomal-rearranged samples had complete/near-complete HPV genome coverage, lacked HPV-human junctions, and contained at least one HPV-HPV junction. 3) Integrated-mixed samples had at least one HPV-human junction and complete/near-complete HPV genome coverage, consistent with coexistence of integrated and episomal HPV DNA. 4) Integrated-clonal samples had at least one HPV-human junction and HPV genome coverage below 97%, consistent with loss of viral sequence between integration sites as well as absence of full-length episomal HPV DNA.

## Junction Confirmation by qPCR

qPCR was performed on cfDNA extracted from plasma and genomic DNA from FFPE tissue. Custom primers targeting selected HPV-human integration junctions and HPV-HPV rearrangement sites were synthesized by Integrated DNA Technologies (IDT); primer sequences are listed in Supplementary Table S1. PCR was performed using the Power SYBR™ Green PCR Master Mix (Applied Biosystems) on a QuantStudio™ 6 Flex Real-Time PCR System (Applied Biosystems). cfDNA from an HPV+HNC plasma lacking the respective integration site was used as a negative control. *GAPDH* was used as a reference gene and 2 ΔΔCt method was used for relative quantification. Junctions were classified as not detected (N.D) if amplification was undetermined or occurred at Ct ≥ 37, which was considered background based on negative-control reactions.

## Orthogonal Junction Detection by Second HPV Whole Genome Sequencing Hybrid Capture Assay

We performed an orthogonal confirmation of the junctions detected by HPV-DeepSeek physical state classification using custom-designed hybrid capture probes developed by an alternative vendor (Twist Biosciences). Hybrid-capture enrichment of 50 HPV genotypes was performed using this probe set (TWI), and sample processing, library preparation, sequencing, and downstream analysis were otherwise carried out according to the described HPV-DeepSeek workflow. For each sample, 2 technical replicates derived from the same cfDNA extraction using TWI probes were analyzed independently and compared with the corresponding HPV-DeepSeek data. Junction calls were considered concordant when breakpoint coordinates matched within ±1 bp. For viral-human junctions, concordance required the same human chromosome. In HPV-HPV rearrangements, the forward and reverse orientations were treated as equivalent.

## Junction Exploration by Long-read Sequencing

Genomic DNA was extracted from fresh frozen tumor tissue using the Monarch High-Molecular-Weight DNA Extraction Kit (New England Biolabs). DNA was sheared to a target fragment size of 15-18 kb using Covaris g-TUBEs (Covaris). Libraries were prepared from 500 ng input DNA using the PacBio SMRTbell Prep Kit 3.0 and sequenced on the PacBio Revio system (Pacific Biosciences) with SPRQ chemistry following the manufacturer’s protocols, targeting ∼30X sequencing depth across the human genome. HiFi reads were aligned with pbmm2 v1.17 to the same custom reference genome used by HPV-DeepSeek. Junctions detected in plasma cfDNA using HPV-DeepSeek were assessed in tumor tissue from aligned PacBio HiFi reads using the same breakpoint-calling workflow. A plasma junction was considered concordant if its representative junction matched a representative junction in the paired tumor tissue within the corresponding clustering tolerance (± 100 bp for HPV-human integrations; ± 20 bp for HPV-HPV rearrangements).

## Survival Analysis

Patient follow-up and outcomes were tabulated using the electronic medical record. Kaplan-Meier survival analyses using a log-rank test and Cox proportional hazard models were performed using RStudio version 2024.12.1. For all statistical analyses, progression-free survival (PFS) and overall survival (OS) were analyzed across broad viral genome physical state classifications, i.e., episomal (episomal-only and episomal-rearranged) vs integrated (integrated-mixed, integrated-clonal) viral states. Cox proportional hazard analysis also incorporated age into the models. A threshold of p < 0.05 was considered significant for all analyses.

## Results

### Cohort Clinical and Genomic Demographics

A total of 235 patients diagnosed with HPV-associated cancer were included in the study cohort, including 195 male patients (83%) and 40 female patients (17%), with a median age of 61 years (range: 36-84). The most common tumor primary site was base of tongue (N = 102) with eight additional anatomic sites included (Figure 1A, Table 1). The most frequently detected HPV genotype in the cohort was HPV16 (N = 204), followed by a diverse array of eight other high risk genotypes: HPV33 (N = 10), HPV35 (N = 6), HPV45 (N = 5), HPV18 (N = 3), HPV56 (N = 3), and HPV67, HPV69, and HPV70 (N = 1 per genotype) (Table 1). Among patient samples harboring HPV16 and for which sublineage analysis was feasible (N = 184), the most commonly detected sublineage was HPV16 A1 (N = 131), followed by A2 (N = 27).

**Figure 1.**
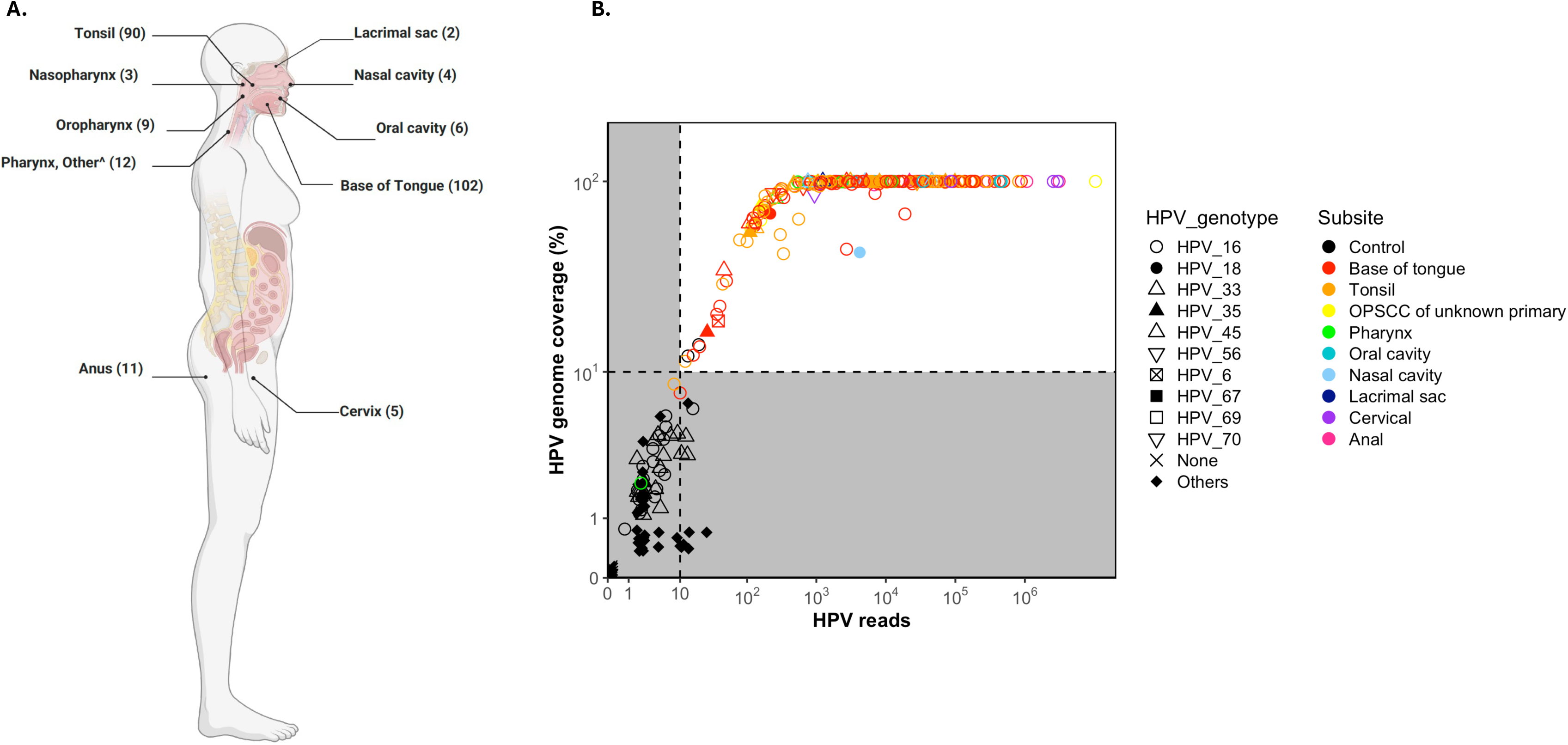
Tumor anatomic locations and diagnostic accuracy of HPV-DeepSeek. (A) Representation of the distribution of tumor primary sites in the study cohort (N = 235). (B) Curve demonstrating HPV detection by NGS using HPV-DeepSeek, focusing on the 2 key metrics for robust identification of HPV in plasma samples. The Y-axis represents percent coverage of the HPV genome, and the X-axis represents the number of supporting reads detected that align to the HPV genome. Samples meeting both thresholds of 10% HPV genome coverage and 10 supportive HPV reads (dashed lines) are considered true positive, represented here by the upper-right quadrant with a white background. Colors represent the anatomic site of the tumor with black representing controls. Shape represents the HPV genotype.

**Table 1.** Cohort Characteristics. The clinical characteristics of the cohort (N = 235) are presented, including median patient age, patient sexes, number of tumors arising in each of 9 primary anatomic sites, and the breakdown of the 10 HPV genotypes detected by HPV-DeepSeek.

|  |  |
| --- | --- |
| **Median age (range)** | 61 years (36-84) |
| **Sex** |  |
| Male | 195 (83%) |
| Female | 40 (17%) |
| **Primary Site** |  |
| Base of Tongue | 102 |
| Tonsil | 90 |
| Pharynx, Other^ | 12 |
| Anus | 11 |
| Oral Cavity | 6 |
| Cervix | 5 |
| Nasal Cavity | 4 |
| Nasopharynx | 3 |
| Lacrimal sac | 2 |
| **HPV Genotypes** |  |
| HPV16 | 204 |
| HPV33 | 10 |
| HPV35 | 6 |
| HPV45 | 5 |
| HPV18 | 3 |
| HPV56 | 3 |
| HPV58 | 1 |
| HPV67 | 1 |
| HPV69 | 1 |
| HPV70 | 1 |

^includes pharyngeal wall, hypopharynx, and cervical lymph node metastases of presumed oropharyngeal origin without known primary

### HPV-DeepSeek Has High Diagnostic Accuracy Across HPV+ Cancers

In the 235 patients diagnosed with HPV+ cancers, HPV-DeepSeek detected a mean of 133,337 reads per sample (median = 4986, range = 2-10,339,813) and a mean coverage of 90% of the HPV genome (median > 99%, range = 2-100%). Across the case-control cohort (N = 447 total) there were 3 false negatives and 2 false positives, yielding 98.7% sensitivity, 99.1% specificity, 99.1% positive predictive value (PPV), 98.6% negative predictive value (NPV), and a 0.98 Youden index (Figure 1B). The 3 false negative samples were all HPV+ oropharynx cancer and were HPV genotype 16 by tissue-based testing, giving a sensitivity of 100% across non-oropharyngeal sites and non-HPV16 genotypes.

### HPV Physical State Can be Classified from the Blood Using HPV-DeepSeek

181 samples could be analyzed for HPV physical state with HPV-SIGNAL based on read thresholds (Methods). Four unique viral genomic patterns of physical state were identified. The first 2 patterns represented episomal varieties of the viral genome, including episomal-only (N = 69, 38%), (Figure 2A) and episomal-rearranged (N = 48, 27%) (Figures 2B). The episomal-rearranged pattern had only HPV-HPV genomic junctions, and no viral-human genomic junctions detected. The final 2 patterns represented viral integration into the human genome, including integrated-mixed (N = 55, 30%) (Figure 2C) and integrated-clonal (N = 9, 5%) (Figure 2D). Among specimens harboring HPV16, the most common genotype of the cohort, the proportions of episomal and integrated viral genome patterns mirrored those of the overall cohort, with 60 (37.5%) episomal-only, 44 (27.5%) episomal-rearranged, 50 (31%) integrated-mixed, and 6 (4%) clonal-integrated identified (Table 2).

**Figure 2.**
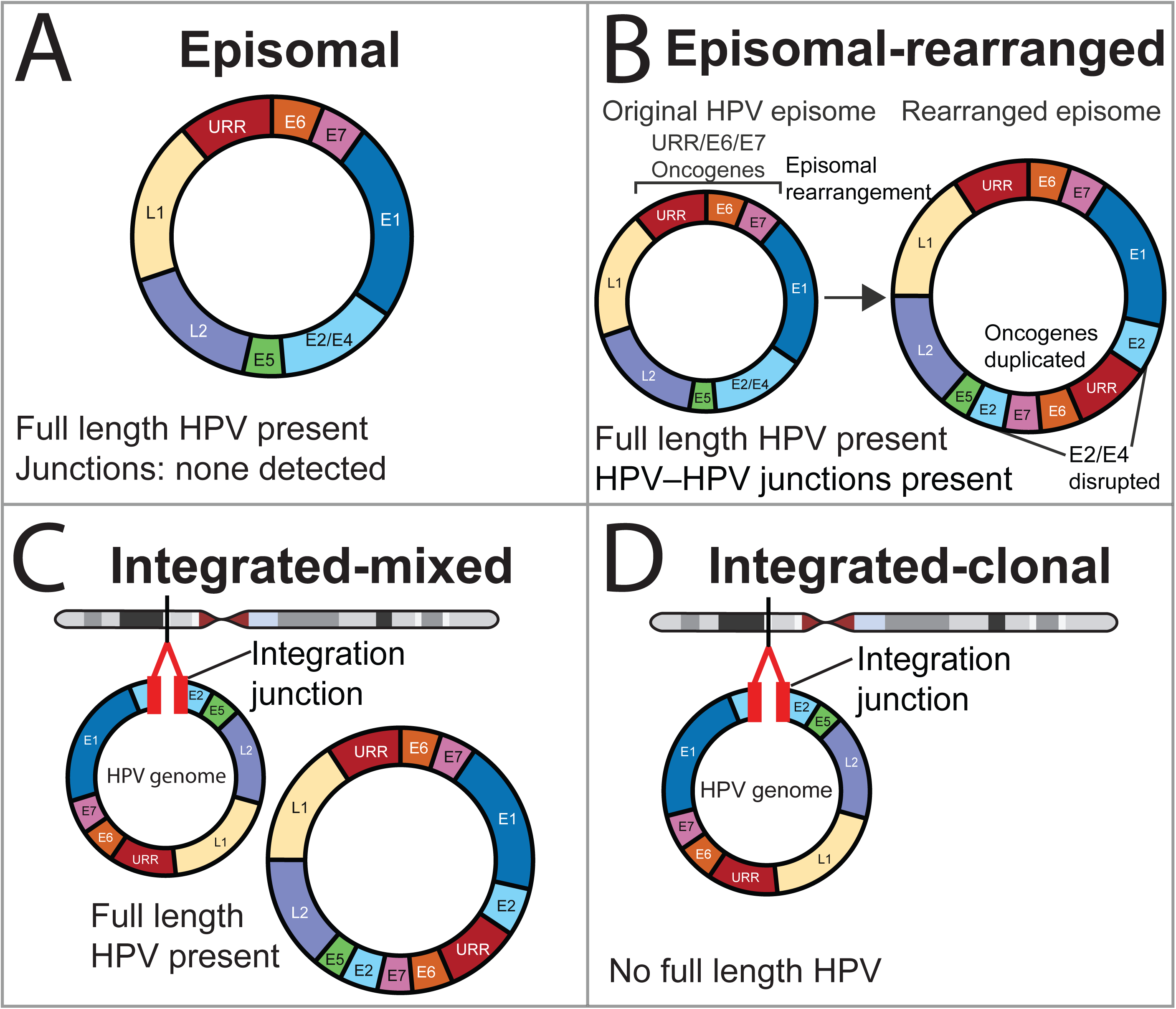
Schematic of HPV16 physical state classification. Colors denote HPV16 genome features (URR, E6, E7, E1, E2/E4, E5, L2, L1), and bright red indicates the HPV-human integration junction. URR, upstream regulatory region; E, early genes; L, late genes. (A) Episomal: full-length circular HPV16 genome is detected, with no HPV-human integration junctions and no HPV-HPV rearrangement junctions. (B) Episomal-rearranged: full-length HPV16 genome is detected together with HPV-HPV junctions consistent with HPV genome rearrangement; the example depicts duplication of the URR/E6/E7 oncogene cassette with disruption of the E2/E4 region, leading to increased genome length (larger size on right in schematic). (C) Integrated-mixed: HPV-human junction reads indicate integration, while concurrent full-length/near-complete HPV16 coverage and HPV-HPV junctions support the presence of episomal HPV molecules in addition to an integrated HPV fragment. (D) Integrated-clonal: a dominant truncated HPV-human integration structure is detected without evidence of a full-length episomal HPV genome.

**Table 2.** HPV Genome Physical State. In this table, the HPV genome physical state, as classified by HPV-SIGNAL following viral genome detection by HPV-DeepSeek, is presented for 181 patients. A total of 4 distinct physical state patterns of the viral genome are identified, including episomal, episomal-rearranged, integrated-mixed, and integrated-clonal. The number of instances of each pattern, as well as a breakdown of how frequently each pattern is seen per HPV genotype is presented.

|  |  |  |  |  |
| --- | --- | --- | --- | --- |
| HPV16 | 60 (37.5%) | 44 (27.5%) | 50 (31%) | 6 (4%) |
| HPV18 | 0 | 0 | 0 | 2 (100%) |
| HPV33 | 6 (100%) | 0 | 0 | 0 |
| HPV35 | 2 (50%) | 1 (25%) | 1 (25%) | 0 |
| HPV45 | 0 | 0 | 3 (100%) | 0 |
| HPV56 | 0 | 2 (100%) | 0 | 0 |
| HPV58 | 0 | 0 | 1 (100%) | 0 |
| HPV67 | 0 | 1 (100%) | 0 | 0 |
| HPV69 | 1 (100%) | 0 | 0 | 0 |
| HPV70 | 0 | 0 | 0 | 1 (100%) |

### Blood-based HPV Physical State Classification by HPV-SIGNAL is Concordant Across Blood and Tissue-based Orthogonal Measures

Representative viral integration and breakpoints were orthogonally confirmed by qPCR in paired tumor tissue and plasma, a second custom HPV hybrid capture NGS assay in plasma, and long-read NGS in paired tumor tissue (Figure 3).

**Figure 3.**
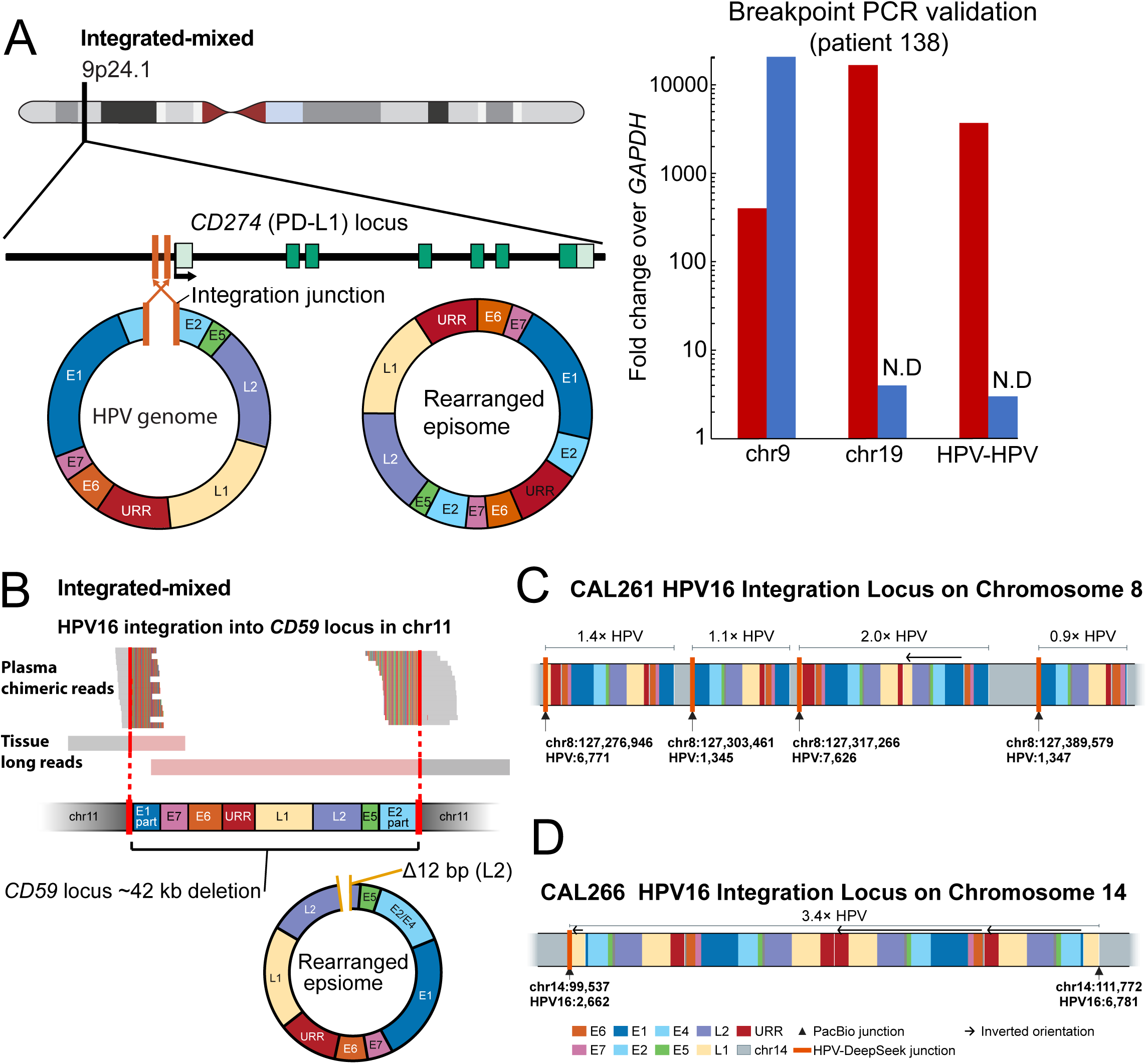
qPCR and long-read sequencing elucidate distinct integrated-mixed and integrated-clonal HPV16 structures detected in plasma cfDNA. (A) Integrated-mixed HPV16 state in patient 138. Plasma cfDNA sequencing identified HPV-human integration at 9p24.1 at the *CD274* (PD-L1) locus together with evidence of rearranged episomal HPV16. The ideogram and locus zoom show the integration site. qPCR assays of three junctions detected by HPV-DeepSeek confirmed the HPV-human chromosome 9 integration site in both plasma (red) and matched FFPE tissue (blue), while confirming the HPV-human chromosome 19 and HPV-HPV integration sites in plasma cfDNA only (red). Signal is normalized to *GAPDH* and displayed as fold change on a log scale. Circular models depict the inferred HPV16 structures. N.D., not detected. (B) Integrated-mixed HPV16 state in patient CAL259. Plasma cfDNA sequencing identified HPV-human junction reads at the *CD59* locus on chromosome 11 together with full-length HPV16 coverage and HPV-HPV junctions, the latter consistent with a rearranged episome containing a 12 bp deletion in *L2*. Tissue long-read sequencing supported a model in which a single ∼5.9 kb HPV16 fragment is integrated in inverted orientation, with a concurrent ∼42 kb deletion spanning *CD59*. The integration disrupts viral *E1/E2* while retaining *URR, E6, E7, E5, L2*, and *L1*. (C) HPV16 integration concatemer at chr8q24.21 in CAL261. Linear schematic of the reconstructed integration locus. Grey blocks indicate retained chr8 segments and colored ribbons indicate HPV16 inserts. Brackets denote total HPV16 content per insert in HPV genome equivalents. Integration junctions were identified by HPV-DeepSeek in plasma cfDNA (orange bars) and by long-read sequencing in matched tissue (black triangles). One insert contains an inverted duplication (left-pointing arrow). Chr8 segments are compressed relative to HPV16, and horizontal distances are not to scale. (D) Palindromic HPV16 integration concatemer at chromosome 14 in CAL266. Linear schematic of the reconstructed integration locus. Grey blocks indicate retained chromosome 14 segments and colored ribbons indicate HPV16 inserts. Brackets denote total HPV16 content in HPV genome equivalents. Black triangles mark integration junctions identified by long-read sequencing, and the orange bar indicates the junction independently detected by HPV-DeepSeek in plasma cfDNA. The concatemer has a palindromic structure, with forward-oriented HPV16 copies on one side and inverted copies (left-pointing arrows) on the other. Chromosome 14 segments are compressed relative to HPV16, and horizontal distances are not to scale. Colors denote HPV16 genome regions (*URR, E6, E7, E1, E2/E4, E5, L2, L1*), and orange bars indicate HPV-human integration junctions. *URR*, upstream regulatory region; *E*, early genes; *L*, late genes.

qPCR was used to orthogonally confirm 3 events detected by HPV-DeepSeek in the plasma of patient 138: 1) an HPV-human junction associated with *CD274* (coding for PD-L1) on chromosome 9, 2) an HPV-human junction on chromosome 19, and 3) an HPV-HPV junction representing a viral genome rearrangement (Figure 3A). The junction associated with *CD274* on chromosome 9 was detected in both plasma cfDNA and matched tumor tissue DNA. The chromosome 19 junction and the HPV-HPV rearrangement were confirmed by qPCR in plasma, but not in paired tissue, further highlighting the strengths of blood-based genomic feature detection to overcome intratumoral heterogeneity.

We used long-read sequencing on matched fresh frozen tumor samples to further elucidate the HPV integration and rearrangement events detected by HPV-DeepSeek in plasma from 3 patients classified by HPV-SIGNAL as integrated-mixed. In CAL259, plasma cfDNA junction reads detected by HPV-DeepSeek identified HPV-human junction reads at the *CD59* locus on chromosome 11 together with full-length HPV16 coverage and HPV-HPV junctions, the latter consistent with a rearranged episome containing a 12 bp deletion in *L2* (Figure 3B). Long-read sequencing confirmed and further elucidated the event in tissue, showing a ∼5.9 kb truncated HPV16 fragment integrated in inverted orientation with a concurrent ∼42 kb deletion spanning *CD59*. The integration disrupts viral *E1/E2* while retaining *URR, E6, E7, E5, L2*, and *L1* (Supplementary Figure S1A).

In CAL261, long-read sequencing identified four HPV16 integration junctions within a ∼113 kb region of chr8q24.21, indicating a multimeric HPV16-human concatemer rather than a single insertion event (Figure 3C). Chimeric reads spanning adjacent junctions showed that HPV16 inserts ranging from 0.9 to 2.0 HPV16 genome equivalents alternate with chromosome 8 segments. One insert contained an inverted duplication of the *E1-L1* region. Three of the four junctions were detected in plasma cfDNA by HPV-DeepSeek with the long-read evidence supporting this reconstruction shown in Supplementary Figure S1B-S1C.

In CAL266, long-read sequencing identified two HPV16 integration junctions on chromosome 14, defining a ∼12.2 kb host deletion. Chimeric reads resolved a palindromic concatemer containing 26,716 bp of HPV16 sequence (3.4 genome equivalents), with HPV16 copies in forward orientation on one side of the structure and inverted orientation on the other. Short inverted *L1* fragments (∼207 bp) bridged both virus-host junctions. One junction was supported by six long reads and detected in plasma cfDNA by HPV-DeepSeek, whereas the second was supported by a single read and was not detected in plasma (Figure 3D, Supplementary Figure S1D). Together, these long-reads confirmed and further elucidated plasma-detected breakpoints and highlight truncated, rearranged, and multimeric HPV16 configurations that could not be resolved by short-read sequencing alone.

We next designed an independent HPV whole-genome hybrid-capture probe panel using a different vendor, with a unique, non-overlapping probe set (TWI), and repeated HPV WGS in 5 plasma samples. HPV-human integration junctions showed high concordance between the orthogonal assay and HPV-DeepSeek, with the majority of junctions shared across approaches for each sample (Figure 4A). We further found high concordance across technical replicates, confirming reproducibility (Figure 4A). HPV-HPV rearrangement junctions assessed by the orthogonal assay also showed cross-platform concordance, though with a larger fraction of probe-design-specific junctions (Figure 4B), likely reflecting differences in viral probe tiling density. A tolerance sweep up to ±1,000 bp rescued no additional matches beyond the ±1 bp threshold, confirming that shared breakpoints are exact positional matches and that unmatched junctions are genuinely probe-design-specific. These results demonstrate that HPV junction breakpoints detected from plasma cfDNA are reproducible across independent capture platforms.

**Figure 4.**
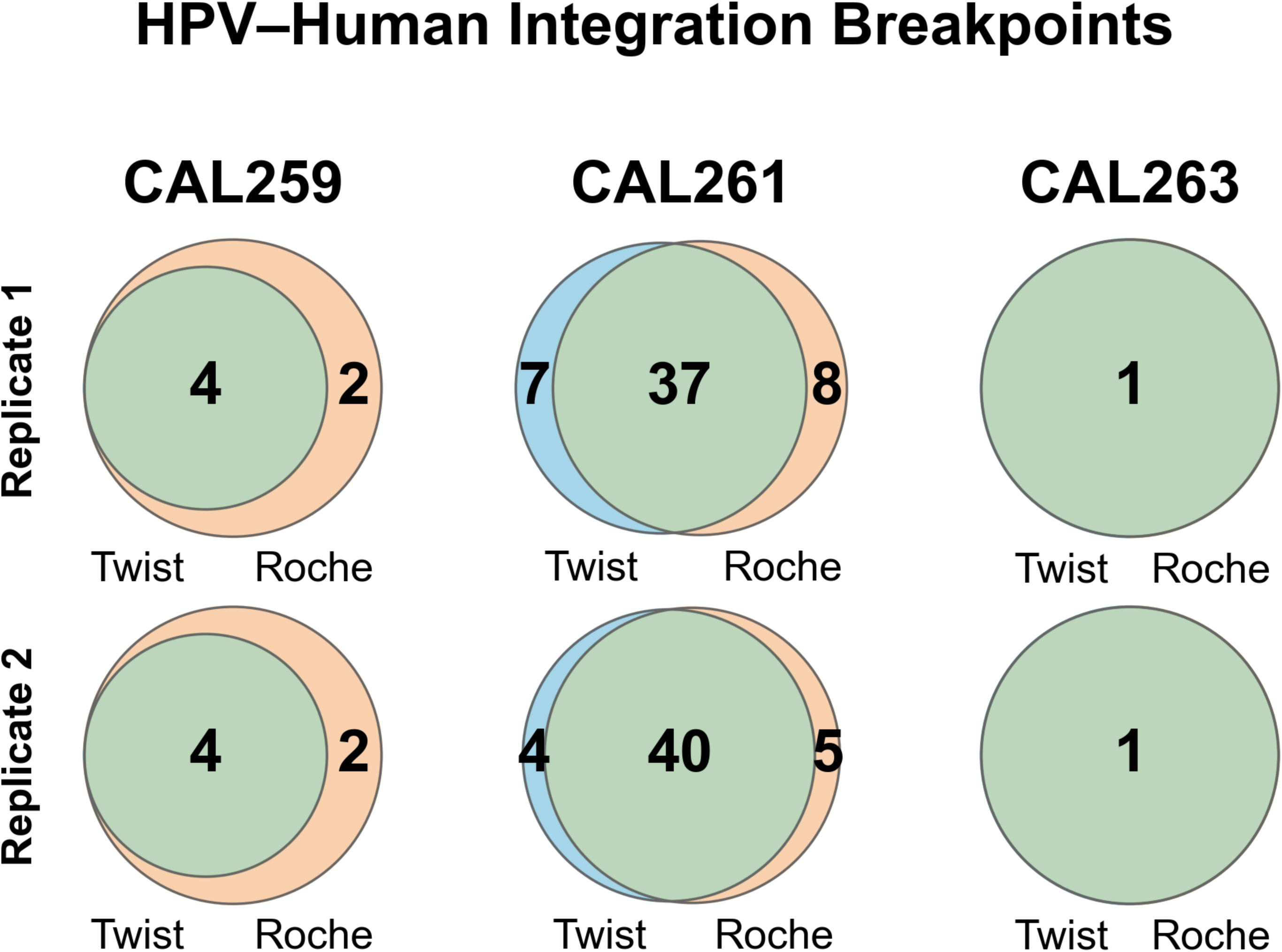
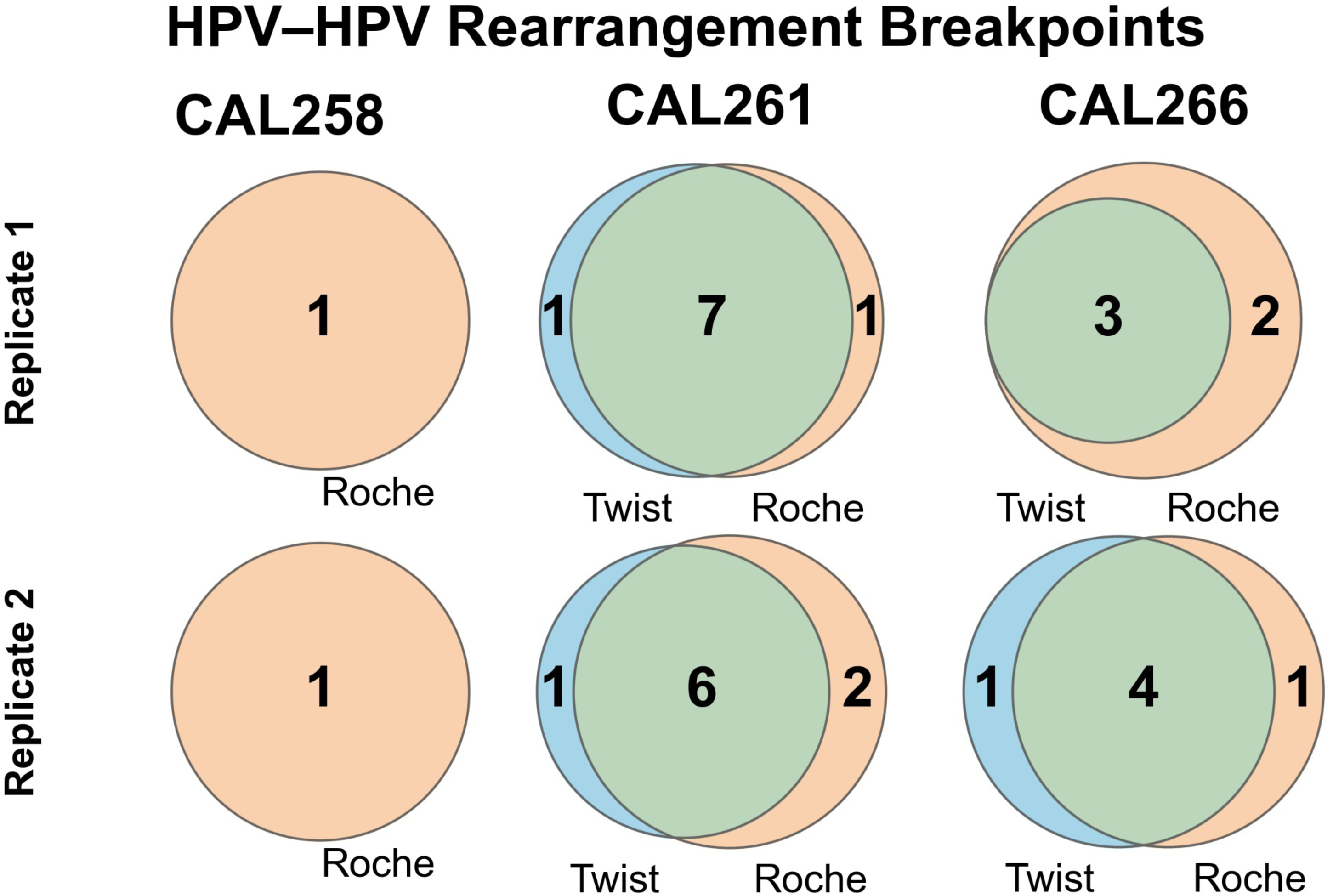
Technical replicate concordance of plasma cfDNA breakpoints across capture probe designs. Venn diagrams showing overlap (green) of junctions detected by Twist (blue) and HPV-DeepSeek (Roche, orange) capture probes for each plasma sample. Columns represent individual patient samples; rows correspond to independent technical replicates, i.e., Replicate 1 and Replicate 2, from the same cfDNA extraction. Numbers indicate junction counts unique to each probe design or shared between both. Junctions were required to have ≥ 6 chimeric supporting reads. Junctions mapping to the HPV genome circular junction (±150 bp from genome termini) were excluded. Samples with no breakpoints of the respective type in either replicate were omitted. (A) HPV-human integration breakpoints were matched at ±1 bp tolerance on human genomic position. (B) HPV-HPV rearrangement breakpoints were matched at ±1 bp tolerance on both junction positions, accounting for forward and reverse orientation.

## Survival Analysis

Oncologic outcomes were assessed for 180 patients with blood collected prior to treatment initiation. The mean follow-ups were 46.9 months (range: 0.8-143.9) for OS, and 39.0 months (range: 0.8-141.6) for PFS. 116 patients were classified as having an episomal viral state (episomal only and episomal-rearranged), and 64 patients were classified as having an integrated viral state (integrated-clonal and integrated-mixed). Consistent with prior tissue-based studies, Kaplan-Meier analysis showed that patients with an integrated state had significantly worse PFS (p = 0.00036; Figure 5A) and OS (p = 0.017; Figure 5B) than those with an episomal state. In Cox proportional hazards models adjusting for age, integrated HPV was associated with significantly worse PFS (HR 3.28, 95% CI 1.63-6.58, p = 0.00084) and OS (HR 2.98, 95% CI 1.16-7.64, p = 0.023) than episomal HPV.

**Figure 5.**
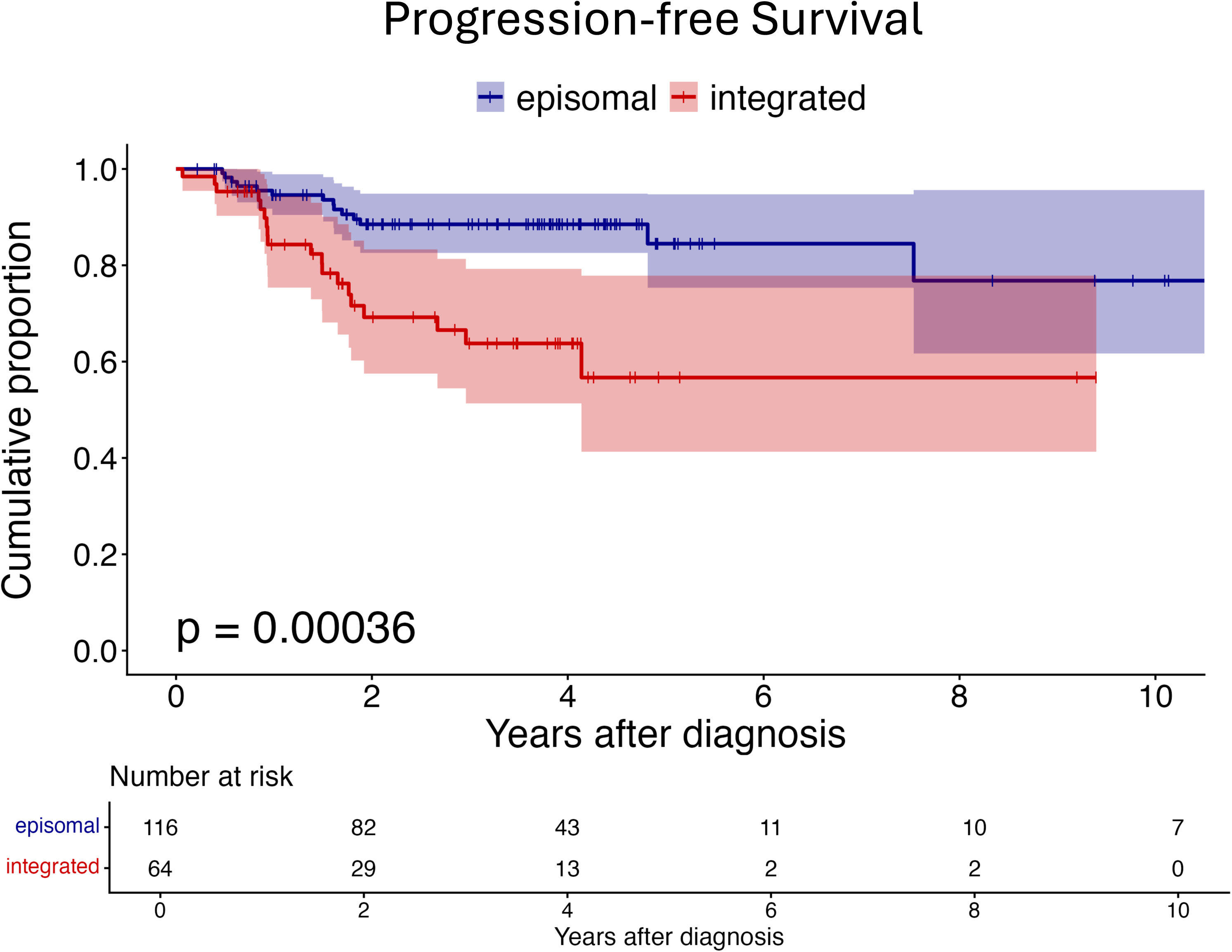
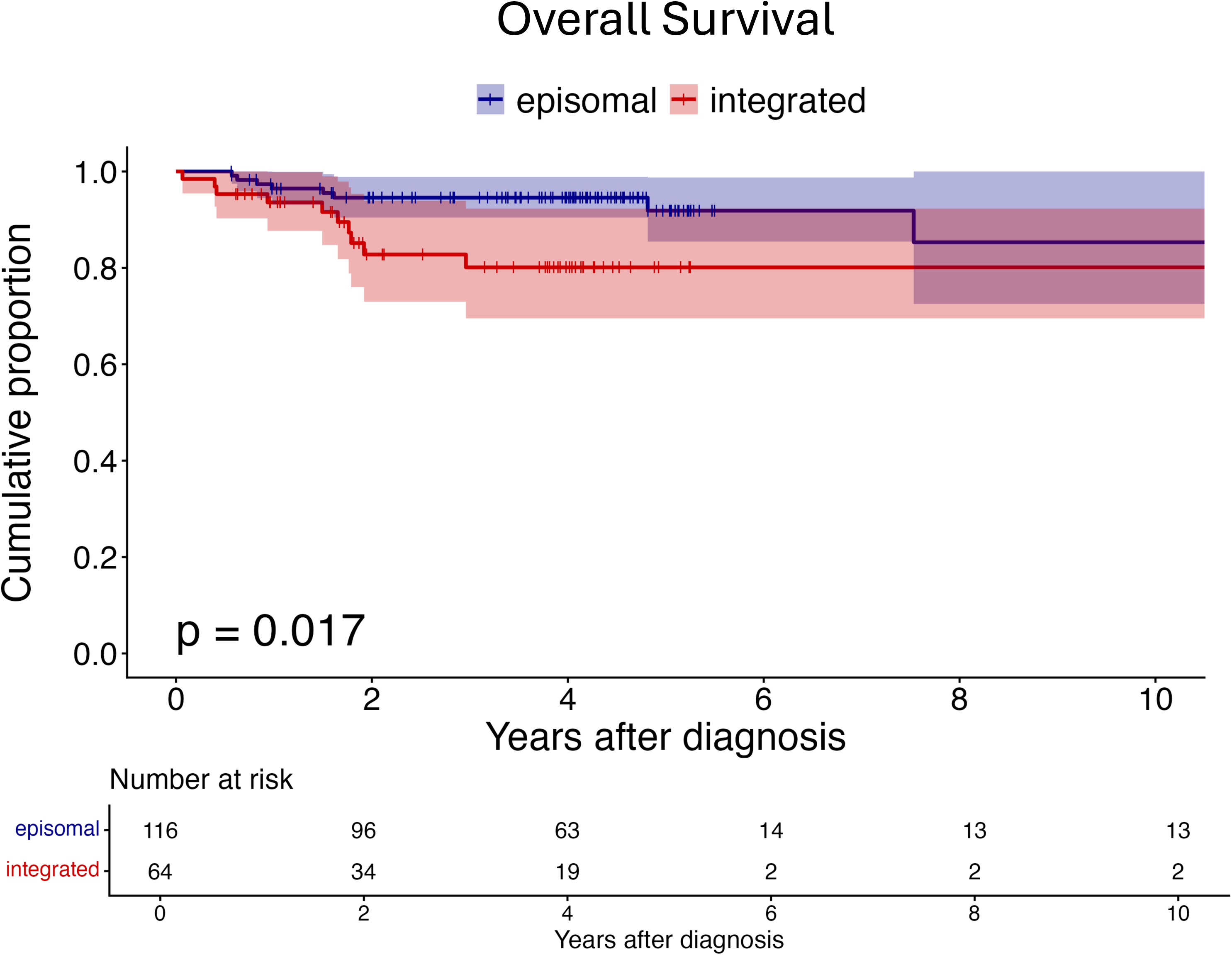
Kaplan-Meier survival analysis with patients stratified by viral genome physical state. (A) Patients with integrated virus (red) had significantly worse progression-free survival (p = 0.00036) than patients with tumors harboring episomal viral genomes (blue). (B) Patients with integrated virus (red) had significantly worse overall survival (p = 0.017) than patients with tumors harboring episomal viral genomes (blue).

## Discussion

Despite the known prognostic value of ctHPVDNA detection in HPV+ cancer surveillance, current ctHPVDNA assays have limited diagnostic accuracy and prognostic utility at the time of cancer diagnosis^3,5–23^. HPV genome physical state is a known prognostic biomarker across HPV+ cancers when detected in tissue biopsies^5,24–27^. In this study we demonstrate the high diagnostic accuracy and prognostic utility of a multi-feature whole-genome sequencing liquid biopsy, across a diverse cohort of HPV+ cancers, finding 99% sensitivity and specificity and enabling the non-invasive classification of HPV physical states directly from plasma using a novel computational classifier, HPV-SIGNAL. Critically, we established that the detection of integrated viral states in the blood is a strong predictor of poor clinical outcomes as measured by decreased PFS and OS. Together, these results support the potential of blood-based HPV physical state profiling as an independent prognostic tool capable of informing risk stratification at cancer diagnosis.

Achieving high diagnostic accuracy at the time of initial presentation of HPV+ cancer is paramount for the downstream clinical utility of ctHPVDNA assays. Robust baseline detection establishes confidence in longitudinal post-treatment surveillance, enables patient accrual for clinical trials requiring detectable ctHPVDNA at diagnosis, and strengthens the assay’s value as a confirmatory diagnostic tool. Building on our previous work demonstrating the high diagnostic accuracy of HPV WGS in HPV+ oropharyngeal cancer, we extend these findings here to non-oropharyngeal cancers, achieving 100% (31/31) sensitivity outside the oropharynx. Importantly, the overall sensitivity and specificity (99%), as well as non-oropharyngeal performance, significantly exceed the metrics reported in the current literature for commercially available ctHPVDNA detection assays^5,9^.

We previously demonstrated the technical capability to detect viral-human junction reads using HPV-DeepSeek, mapping them to specific human and viral genomic locations^7^. Here, we extend these findings by developing a computational classifier, HPV-SIGNAL, that synthesizes multiple lines of evidence, integrating data not only from viral-human junctions, but also from HPV-HPV structural rearrangements, overall HPV genome coverage, and thresholds for required read quantities. Application of HPV-SIGNAL enabled the confident assignment of samples into one of four mutually exclusive viral physical states: episomal, episomal-rearranged, integrated-mixed, and integrated-clonal. To validate and further elucidate of these classifications, we employed three approaches. First, we performed PCR on paired plasma and tissue biopsies. PCR analysis of paired plasma definitively confirmed the presence of HPV-SIGNAL-detected events. Notably, several events identified in circulation were absent in corresponding tissue biopsies, likely reflecting the inherent advantage of liquid biopsies in capturing the full spectrum of intratumoral genomic heterogeneity compared to spatially limited tissue sampling³ . Second, the use of an independent HPV whole-genome hybrid-capture probe panel demonstrated high concordance, with minor discrepancies in junction detection attributed to probe tiling density rather than biological discordance—a finding that underscores the impact of probe design on analytical sensitivity. Lastly, long read WGS was used to confirm and further elucidated plasma-detected breakpoints, highlighting truncated, rearranged, and multimeric HPV16 configurations that could not be resolved by short-read sequencing alone.

Most critically, we demonstrate that blood-based HPV genome physical state classification is not only technically feasible but carries significant prognostic value across HPV+ cancers. These findings suggest that a multi-feature HPV WGS liquid biopsy can fulfill a major clinical need by enhancing prognostic stratification prior to the initiation of treatment. In our cohort, the detection of integrated viral states in plasma was a robust predictor of poor oncologic outcomes; specifically, patients with integrated states exhibited significantly worse PFS and OS compared to those with episomal states. Notably, these associations remained statistically significant in Cox proportional hazards models after adjusting for patient age. By enabling non-invasive classification at the time of diagnosis, such an approach could offer a scalable tool for identifying patients who may benefit from intensified or de-intensified treatment. These finding further validate tissue-based studies demonstrating worse outcomes in integrated tumors across HPV+ cancer types, but now offer a clinically implementable approach for determining the status of this biomarker.

The mechanisms by which HPV integration drives a more aggressive phenotype and poorer prognosis remain an area of investigation. HPV integration has been shown to induce focal genomic instability, as integration events are frequently found flanking complex structural variants within the human genome, including copy number variants and large-scale genetic rearrangements. Beyond local disruption, integration can trigger the expression of viral transcripts and drive the overexpression of host oncogenes through insertional mutagenesis and the introduction of viral enhancers.

This study has a number of limitations that warrant consideration. First, although the cohort is diverse, HPV+ oropharynx cancers make up a disproportionate number of cases. Additional cohorts with larger numbers of anogenital cancers should be included in future validation cohorts to confirm that the prognostic significance of blood-based viral physical state classification remains consistent across all HPV+ cancer types. Importantly, HPV+ oropharynx is by far the most common HPV+ cancer in the United States. Second, the HPV-SIGNAL classifier requires a minimum read threshold for confident physical state assignment, which excluded a portion of the total cohort from prognostic analysis. While this ensures high analytical specificity, clinical impact would be restricted to cases meeting classification thresholds. Future refinements of HPV-SIGNAL will be needed to classify samples with very low ctHPVDNA concentrations. Third, although our survival analysis demonstrated a clear prognostic link between integrated states and worse PFS and OS, these findings were established in a retrospective cohort and should be validated in prospective trials to confirm their utility in directing treatment intensification or de-intensification.

In summary, a multi-feature HPV WGS liquid biopsy, HPV-DeepSeek, has high diagnostic accuracy across HPV+ cancers regardless of anatomic site and, empowered by HPV-SIGNAL, classified HPV physical state in plasma, demonstrating prognostic utility for recurrence and survival. These findings broaden the clinical utility of HPV liquid biopsy and suggest HPV liquid biopsy could be used at the time of cancer diagnosis to identify patients for escalated or de-escalated care.

Funding for this work came from NIH/NIDCR R03DE030550 (Daniel L. Faden).

## Data Availability

All data produced in the present work are contained in the manuscript.

