## Supplemental Figures for "Whole Genome HPV Liquid Biopsy for Pan-HPV-Associated Cancer Detection and Viral Physical State Classification"

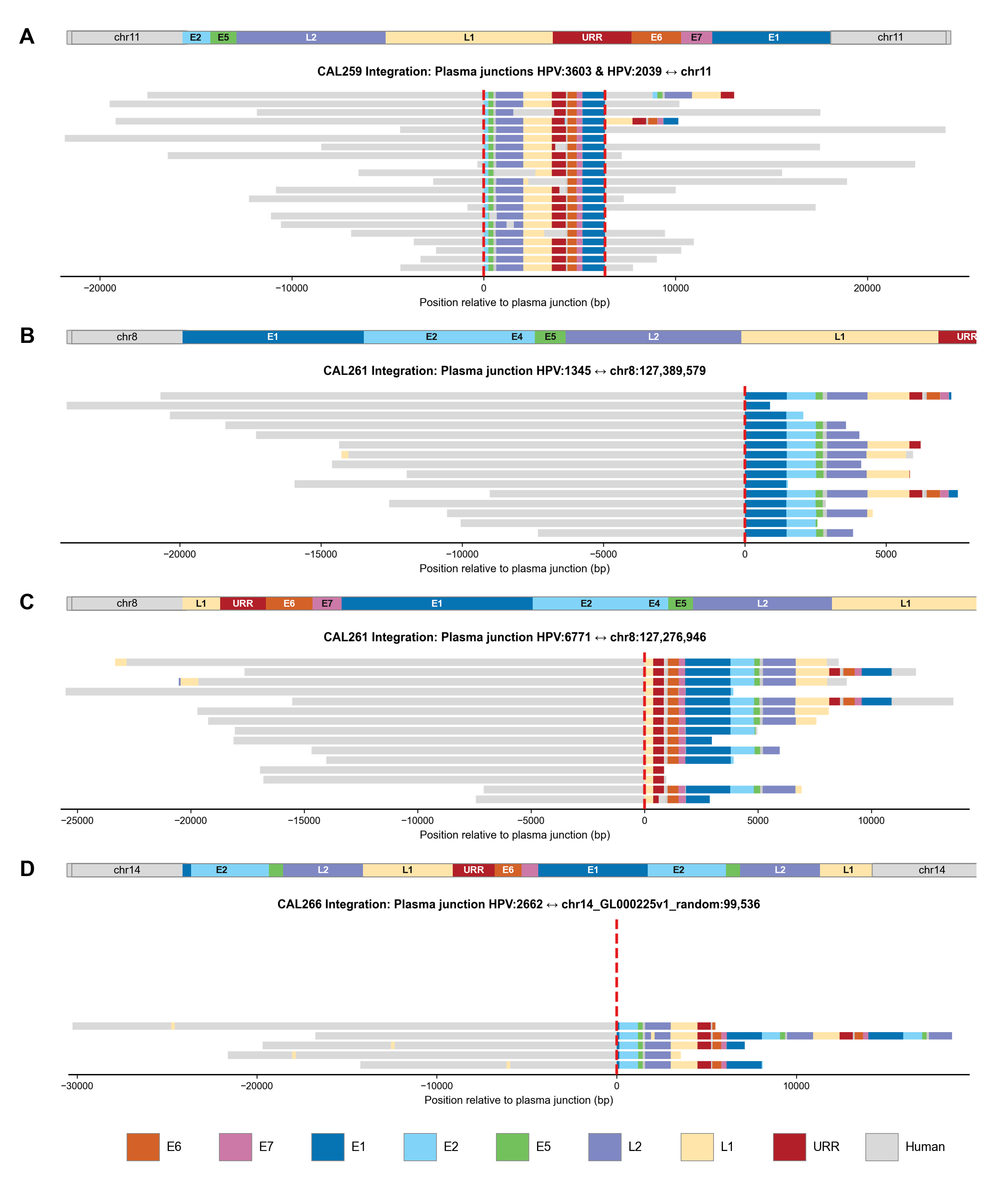
**Supplemental Figure 1**

**Supplemental Figure S1.** Long-read sequencing performed on matched fresh frozen tumor tissue with reads spanning plasma-detected junctions are shown for 3 cases. Each horizontal bar represents a single long read, color-coded by HPV16 gene annotation (E6, E7, E1, E2, E4, E5, L2, L1, URR) or human genomic sequence (gray). The red dashed line marks the plasma cfDNA junction position detected by HPV-DeepSeek. (A) CAL259 shows 2 HPV16-human junctions linking HPV16 positions 3603 and 2039 to separate loci on chromosome 11, consistent with a ~5.9 kb truncated HPV16 fragment integrated at the *CD59* locus. (B) CAL261 shows four HPV16-human junctions linking HPV16 positions 1345, 6771, 2148, and 1129 to nearby loci on chromosome 8, revealing a complex multimeric HPV-human concatemer with fragmented and repeated viral segments rather than a single integration event. (C) CAL266 shows one HPV16-human integration junction linking HPV16 position 2662 to chromosome 14 and one intraviral HPV16 junction linking positions 4618 and 5868, consistent with coexisting chromosomal integration and internal viral rearrangement, including an approximately 1.25 kb deletion across the L2/L1 boundary. Up to 30 representative reads are displayed per junction. (D) CAL266 shows HPV16-human integration on chromosome 14. Five reads show integration breakpoints at HPV16 positions 2662 and 6572, flanked by chromosome 14 (chr14_GL000225v1_random) sequence. The integrated HPV16 segment spans ~11.8 kb, larger than the 7906 bp canonical genome because it contains an approximately 3.9 kb duplicated region between positions 2662 and 6572. Despite this rearrangement, E6, E7, and URR are retained. Only reads containing a clear HPV-human or HPV-HPV transition at the expected breakpoint position, with at least 300 bp of aligned sequence flanking each side of the junction, are shown.
